# A mixed-methods evaluation of the implementation of IOTA-ADNEX ultrasound triage in NHS secondary care ovarian diagnostic one-stop clinics

**DOI:** 10.1101/2025.11.23.25340389

**Authors:** Vivian Do, Helen Crisp, Carole Cummins, Sarada Kannangara, Grisham Smotra, Becky Tarbuck, Omiete Duke, Aamena Salar, Nina Jhita, Vincent Sai, Naresh Rati, Sudha Sundar

**Affiliations:** Pan Birmingham Gynaecological Cancer Centre, Sandwell and West Birmingham NHS Trust, Birmingham, B66 2QT; Department of Cancer and Genomic Sciences, School of Medical Sciences, College of Medicine and Health, University of Birmingham, B15 2TT; Crisp Quality Improvement Consultancy Limited, London SE5 8NJ; Department of Obstetrics and Gynaecology, Walsall Healthcare NHS Trust, Walsall, WS2 9PS; Primary Health Care and Community Services, Modality Partnerships, UK

## Abstract

**Objectives:** Robust evidence supports IOTA-ADNEX ultrasound triage at 10% threshold for ovarian cancer (OC) diagnosis to identify women for referral to tertiary gynaecological cancer centres for further management. The IOTA-ADNEX risk prediction model has superior sensitivity compared to the current standard of care triage, Risk of Malignancy Index (RMI-1), yet NHS adoption is limited. In our survey of British Gynaecological Cancer Society clinicians only 30% (24/79) currently follow an IOTA model, despite 80% (63/79) supporting implementation. We evaluated IOTA-ADNEX implementation within two NHS one-stop clinics (OSC) for suspected OC, examining clinical outcomes alongside implementation barriers and facilitators.

**Methods:** Mixed-methods study conducted across two UK NHS hospitals between June 2023-June 2025. Implementation outcomes were surgical intervention rates comparing IOTA-ADNEX-guided and retrospectively calculated RMI-based management using NICE/RCOG thresholds and patient process metrics. 11 qualitative semi-structured interviews were conducted with NHS staff involved in OSC implementation and thematic analysis performed.

**Results:** Of 334 patients, 42% (139) underwent same-day discharge. Using IOTA-ADNEX at a 10% threshold, 10% (32/334) of patients underwent surgery under the general gynaecology and cancer unit team. In comparison, 30% (94/334) would have undergone surgery under the same teams if RMI-based triage had been used. Five themes identified from qualitative analysis: organisational infrastructure, clinical decision-making, communication and pathway definition, professional collaboration and training support, and patient experience. Key facilitators included dedicated clinical leadership, timely decision-making capabilities and quality assurance sessions. Barriers included lack of standardised post-clinic pathways and insufficient staff communication about pathway changes.

**Conclusions:** IOTA-ADNEX implementation in one-stop clinics offer high same-day discharge rates and reduction in surgical rates compared to RMI triage. To ensure success, implementation should be supported by adequate infrastructure, training, and clear pathways. It requires leadership, comprehensive staff training, and robust communication strategies. These findings provide practical guidance for healthcare systems for wider implementation of IOTA-ADNEX.

**Key messages:** The IOTA-ADNEX ultrasound risk-assessment model to triage adnexal masses has demonstrated superior diagnostic accuracy over RMI which remains the standard tool in UK ovarian cancer pathways, even when applied by certified non-expert sonographers, but implementation within UK NHS pathways remains limited.

This mixed-method evaluation shows IOTA-ADNEX ultrasound triage can be successfully implemented in NHS one-stop clinics, reducing unnecessary benign surgeries compared to RMI and enabling high same-day discharge rates without missing invasive cancers.

Findings on key facilitators and barriers highlight the need for infrastructure, training, and pathway clarity and will inform wider NHS adoption of IOTA-ADNEX.

## Introduction

Ovarian cancer (OC) represents a serious health challenge and remains the 6^th^ most common cancer in the UK.(1) Recent UK statistics are alarming: approximately 30% of women die within their first year after diagnosis and 70% present with advanced-stage diseases.(2) Late-stage presentation dramatically impacts survival chances where five-year survival is only 13% (stage IV), compared to over 90% for stage 1.(1) Diagnosis is often delayed due to vague symptoms including bloating and abdominal pain which can be attributed to less serious conditions. Efforts have been made to enhance diagnostic accuracy, particularly through robust triaging of adnexal (ovarian) masses, which ensures timely diagnosis and referral to the appropriate care setting. UK OC care is delivered through the National Health Service (NHS) using a hub-and-spoke model. Local general hospitals (‘spoke’ sites) conduct initial assessments and manage lower-risk cases via the gynaecological cancer unit team. Patients with suspected or high-risk OC are referred to ‘hub’ centres, where specialised multidisciplinary teams including gynaecological oncologists, are available to offer advanced surgery, cancer treatments, comprehensive peri-operative and psychosocial support.(3–5) Centralising these cases to specialised cancer centres improves patient outcomes, with 10% lower mortality compared to management in non-specialist settings (general hospital).(3) This underscores the importance of accurate triaging to ensure that the right patients reach the right expertise at the right time.

The UK’s National Institute for Health and Care Excellence (NICE) recommends a stepwise referral pathway for suspected OC (supplementary material).(6) Primary care physicians request a CA125 blood test (protein which can be raised in OC) for symptomatic women, followed by a pelvic ultrasound if raised. Abnormal scans prompt urgent hospital referral for patients to receive a definitive diagnosis within 28 days under NHS Faster Diagnosis Standards.(7) In practice, referral patterns often deviate from this pathway, with patients referred with symptoms alone or with partial test results. Hospital doctors calculate the Risk of Malignancy Index (RMI), a score combining ultrasound findings, CA125 levels and menopausal status, to estimate the cancer risk.(6) RMI scores <25 indicates low risk (<3% cancer probability), 25-250 moderate risk (20% cancer probability) and >250 high-risk (70% cancer probability).(8) High-risk patients (RMI≥250) are referred to cancer centres for open surgery by gynae-oncologists, while lower risk patients (RMI<250) receive monitoring or minimally invasive surgery by local gynaecology or cancer unit teams. NICE recommends a threshold of 250, while the Royal College of Obstetricians and Gynaecologists (RCOG) applies 200 for postmenopausal women.(6,8) Both offer comparable diagnostic performance, with sensitivities of 70% and 78% and specificities of 90% and 80%, respectively.(8)

Compared to RMI, the IOTA (International Ovarian Tumour Analysis)-ADNEX (Assessment of Different NEoplasias in the adneXa) model offers a superior diagnostic accuracy (AUC 0.93 vs 0.82) and misses less cancers with a higher sensitivity (96.1% vs 82.9%).(9–11) It is a multiclass ultrasound risk-prediction model which differentiates between a non-cancerous and cancerous mass, and breaks down the cancer risk into borderline tumour, early-stage (stage I), advanced stage (stage II-IV) and secondary metastases (cancer has spread to the ovary from elsewhere). It remains accurate even when used by non-expert, IOTA-certified, quality-assured sonographers.(11) Its adoption is supported by expert consensus and incorporated into British Gynaecology Cancer Society (BGCS) 2024 guidelines.(12,13)

During a single ultrasound appointment, ultrasound examiners apply the IOTA-ADNEX model using a two-step strategy to evaluate the adnexal masses. First examiners assess for specific ultrasound features termed ‘modified benign descriptors’ (BD) to identify masses likely to be benign with a cancer risk of <1%. If not applicable, IOTA-ADNEX score is calculated.(14) Validation in long-term studies shows complication rates <1% for conservatively managed benign masses over two years.(15) Embedding this strategy into one-stop clinics (OSC), where patients receive consultation, ultrasound and management plan in a single visit can streamline care, reduce unnecessary imaging and appointments. The OSC model has proven successful in other NHS gynaecological pathways including endometrial cancer.(16,17)

Evidence on its implementation is limited, our systematic literature search identified one paper addressing current uptake and barriers of IOTA models in clinical practice.(18) In our survey of BGCS clinicians only 30% (24/79) currently follow an IOTA model, despite 80% (63/79) supporting implementation. Successful implementation requires more than clinical validation: it depends on infrastructure, workforce engagement, resources and consideration of patient experience. We explored the clinical impact, facilitators and barriers of implementation in a mixed-methods study.

## Methods

### Study design

This quality improvement study employed a convergent parallel mixed-method design, integrating quantitative and qualitative data to evaluate both clinical impact and implementation experience of the adoption of IOTA-ADNEX two-step strategy within NHS OSC. Quantitative and qualitative components were conducted concurrently, analysed independently, and merged during interpretation to provide a comprehensive understanding of the intervention’s effectiveness and contextual feasibility.

### Quantitative Component

A multicentre, prospective, observational cohort study evaluated symptomatic women attending weekly ovarian OSC at two NHS hospitals from June 2023 (Sandwell and West Birmingham, SWBH, a cancer centre) and October 2023 (Walsall Healthcare NHS Trust, a cancer unit) until June 2025. Clinics offered same-day gynaecological assessment and pelvic ultrasound (transabdominal and transvaginal) by IOTA-certified Level II examiners and reported using standardised IOTA terminology, providing a structured and standardised description of the masses.(19) The IOTA-ADNEX two-step strategy was applied in the same appointment to triage masses, guiding clinical management based on the multi-panel expert consensus (2021) (Figure 1). The IOTA-ADNEX calculator was made available through Professor Dirk Timmerman (now available as a medical device app through the Gynaia website).(20) During early implementation, IOTA Simple Rules (SR) model was used (n=24) due to temporary calculator unavailability. This model classifies adnexal masses as benign (‘B’ rules), malignant (‘M’ rules) or ‘Uncertain’ if both rules apply. Walsall used a single IOTA-trained gynaecologist for scanning and consultation, while SWBH employed IOTA-certified doctors working with trained sonographers.

**Figure 1:**
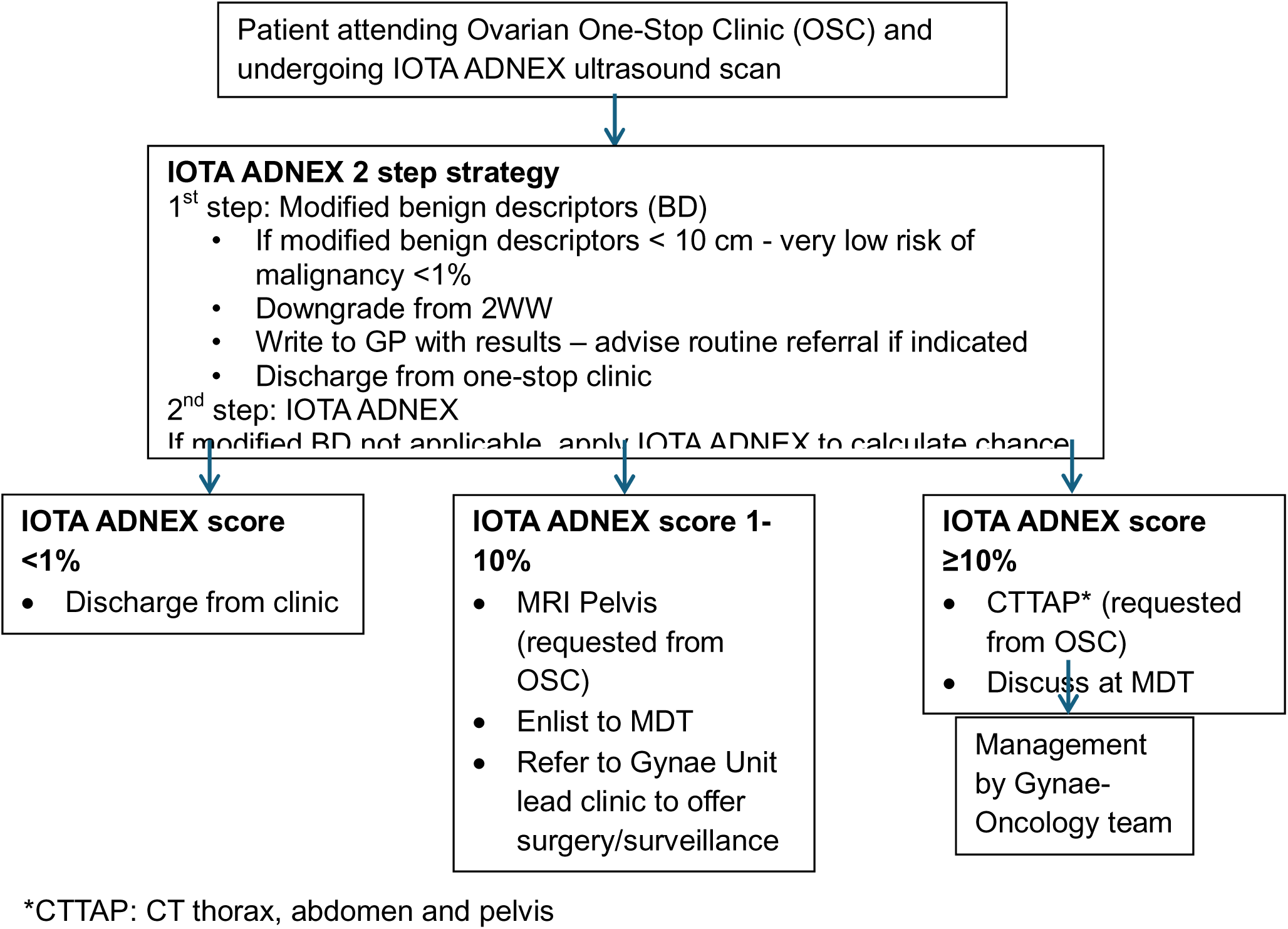
One-stop clinic flowchart used at SWBH (Modified from 2021 ESGO guidance)

Patient data were extracted from electronic health records including demographics, ultrasound findings, triage decisions, surgical outcomes, and discharge timelines by NHS employed clinical research fellows. Data were stored and analysed securely on an Excel spreadsheet on NHS One Drive. Clinical decisions were based on IOTA-ADNEX triaging at 10% threshold applied in real-time, while RMI scores were retrospectively calculated using the formula: U (Ultrasound score) x M (menopausal status) x CA125 (IU/ml).(6)

#### Inclusion criteria

Women referred via the urgent suspected OC pathway from primary or secondary care.

#### Exclusion criteria

Pregnancy and patients who did not attend their appointment.

The primary outcome compared surgical intervention rates based on IOTA-ADNEX-guided decisions and retrospectively calculated RMI-based management using NICE/RCOG thresholds. Secondary outcomes were process metrics: additional imaging requirements and discharge patterns (within three months of referral/same day). Patients not discharged during this period due to ongoing investigation and management were classified as ‘follow-up with surveillance’. Data were analysed using descriptive statistics calculated in Excel. Borderline ovarian tumours were classified within the ovarian tumour category for analysis purposes. In the UK, surgical management of suspected borderline tumours may be undertaken in either cancer unit or cancer centre settings.

### Qualitative Component

This was a prospective, qualitative evaluation study, looking at the facilitators and barriers of IOTA-ADNEX implementation across the two NHS OSC.

11 semi-structured interviews involving 14 purposively sampled NHS staff were conducted throughout the study period (SWBH: June2023, Walsall: October 2023 until June 2025). Participants included gynaecologist, gynae-oncologists, sonographers, clinic and cancer service administrative staff. Virtual interviews (30-35minutes) were conducted via Microsoft Teams/Zoom by an independent Quality Improvement consultant (HC) and a gynae-oncology research fellow based at SWBH (VD). Real-time transcripts were documented on Microsoft Word. Interviews were deemed to have achieved data saturation, as no new themes or insights emerged in the last few interviews. Inductive thematic analysis followed Braun and Clarke’s methodology, with independent coding by HC and VD.(21)

### Ethics

This is an NHS quality improvement project which received local governance approval and audit permissions from Sandwell and West Birmingham NHS Trust (Audit 3111) and Walsall Healthcare NHS Trust, without requiring Health Research Authority ethical review, as it represented evidence-based standard care implementation with no additional patient risks.(13) All data were anonymised to ensure patient confidentiality.

## Results

Over the study period, 340 patients were referred, and 334 patients were seen in the OSC. Six patients were excluded: 2 patients were pregnant and 4 did not attend the appointment (Figure 2). Patient characteristics are summarised in Table 1. 92% (306/334) of patients underwent IOTA ultrasound assessment. Of those who did not undergo IOTA assessment (28/334), most had prior imaging (n=23) and the remaining declined IOTA ultrasound (n=5). 55% (169/306) had previous pelvic ultrasounds before IOTA-ADNEX assessment.

**Figure 2:**
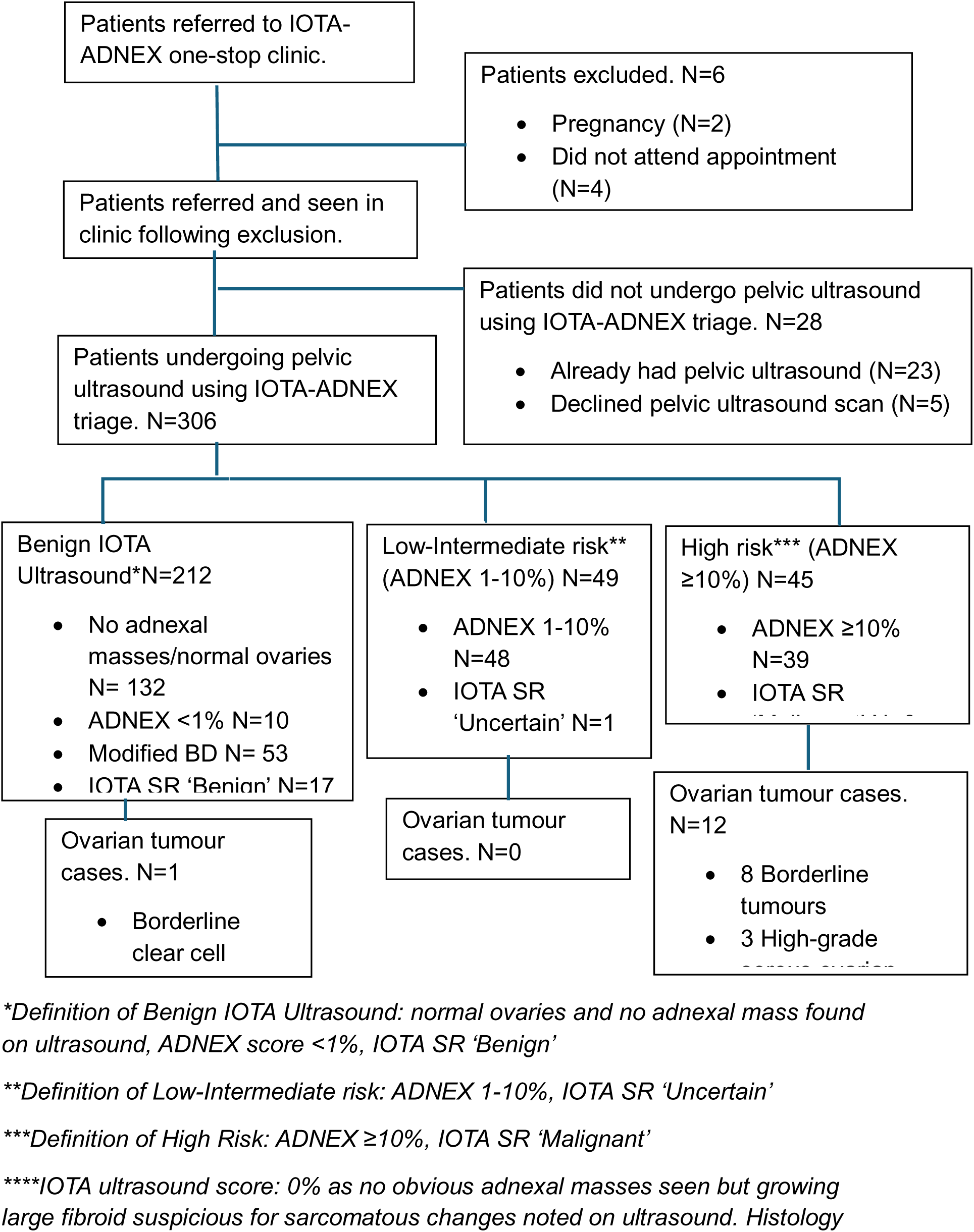
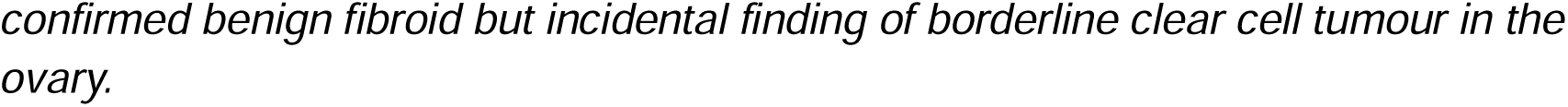
Flowchart of patients included, excluded, undergoing IOTA scans and ovarian tumour outcomes, N (number of patients)

**Table 1:**
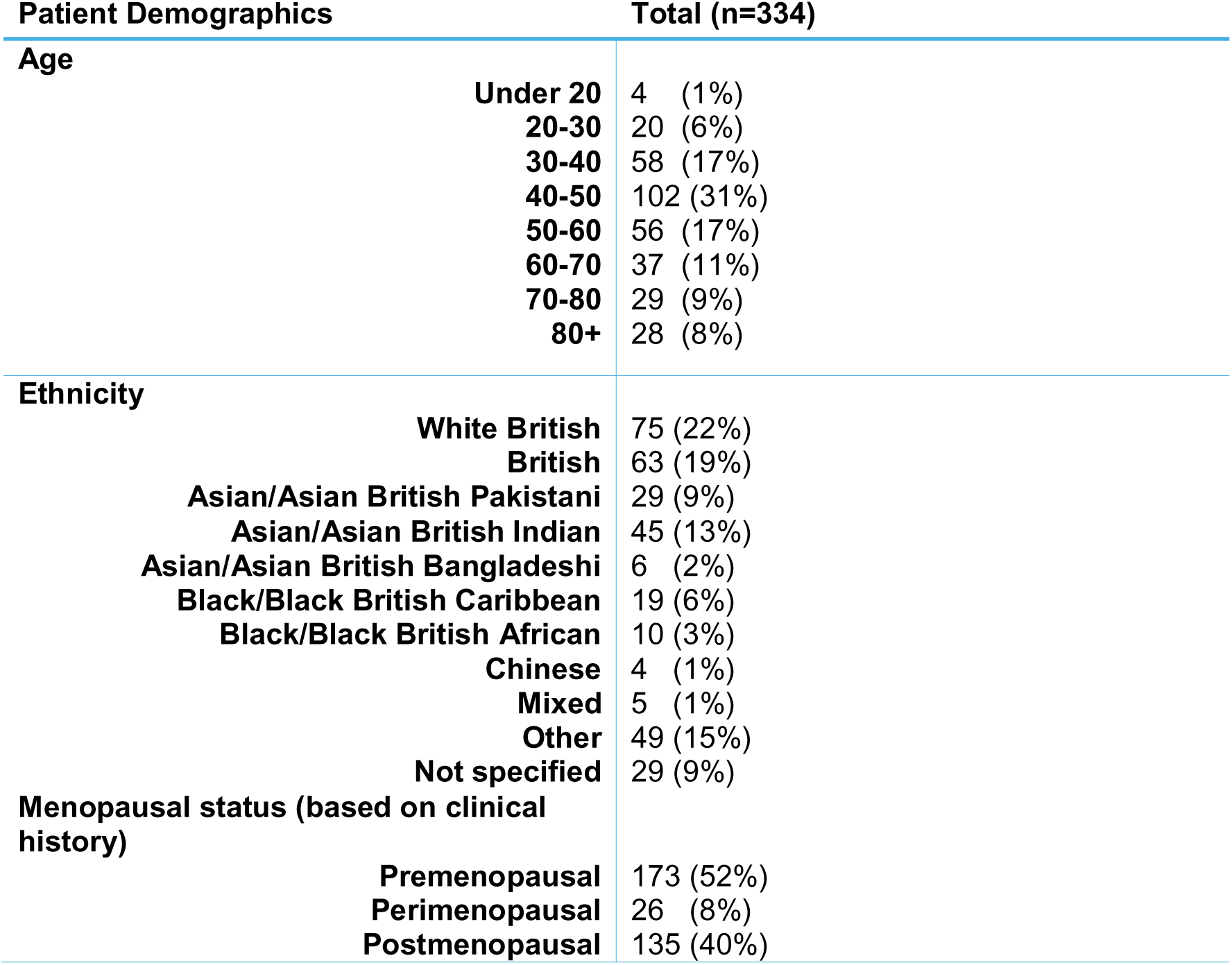
Patient characteristics: Age group, ethnicity and menopausal status.

A total of thirteen histologically confirmed ovarian tumour cases (4%) were diagnosed: 9 borderline tumours (serous borderline n=6, mucinous borderline n=2, clear cell borderline n=1) and 4 invasive OC (high-grade serous n=3, clear cell carcinoma n=1). All invasive OC were identified following the IOTA-ADNEX scan with no missed invasive OC during this study period.

### Benign IOTA findings (n=212, 69%): Ovarian tumour cases n=1

Benign IOTA classification was defined as ultrasound reporting normal ovaries or no obvious adnexal masses (n=132), ADNEX score <1% (n=10), modified BD (n=53) or IOTA SR ‘Benign’ (n=17). 7 underwent surgery and had confirmed benign histology, the remaining 7 patients are awaiting surgery under the general gynaecology team.

One borderline clear cell adenofibroma ovarian tumour was diagnosed histologically. In this case, the IOTA ultrasound was benign with an ADNEX score of 0% as scans did not show any obvious adnexal masses. Patient underwent surgery for suspected sarcomatous changes within a large fibroid.

### Low-Intermediate Risk Group: IOTA-ADNEX -10% (n=49, 16%): Ovarian tumour cases n=0

48 patients were found to have an IOTA-ADNEX score of 1-10% and 1 patient was deemed ‘Uncertain’ using IOTA SR. No cancer cases were diagnosed, and the majority (6/11) of patients had confirmed benign histology, the rest are pending surgery under the general gynaecology team.

### High Risk Group: IOTA-ADNEX ≥10% (n=45, 15%), Ovarian tumour cases n=12

39 patients had an IOTA-ADNEX score ≥10%, 6 with IOTA SR classification ‘Malignant’. Following surgery, benign histology was found in twelve patients, two patients are pending surgery (cancer centre n=1, general gynaecology n=1). One patient has undergone surgery with intraoperative appearances suggestive of disseminated OC and histology is currently pending. This group yielded the highest ovarian tumour detection rate (n=12, 27%). Among patients who underwent surgery with cancer centre there were eight borderline ovarian tumours, three high grade serous OC (stages: 3A1ii, radiologically stage 4B and one un-staged). One clear cell carcinoma was diagnosed following surgery with a cancer unit team.

### Surgical outcomes following IOTA-ADNEX compared with_RMI at 250-threshold

Surgical outcomes following IOTA ultrasound scan and predicted RMI based on 250-thresholds are shown in Table 2. Using IOTA-ADNEX at a 10% threshold, 10% (32/334) of patients underwent surgery under the general gynaecology and cancer unit team. In comparison, 30% (94/334) would have undergone surgery under the same teams if management was based on retrospectively calculated RMI using NICE/RCOG thresholds. 62% (8/13) of diagnosed ovarian tumours had a calculated RMI≥250. The remaining, three borderline ovarian tumours and two invasive cancers (one clear cell OC, one high grade serous OC) had a RMI score <200: these patients would not have been managed in a cancer centre setting had RMI been used.

**Table 2:** Patients undergoing surgery following IOTA scan and predicted RMI triage to surgery based on 250 threshold.

| Menopausal status | Surgical outcomes following IOTA scanning (n=306) |  |  | Total | Predicted triage to surgery in general gynaecology/ cancer unit RMI 25-250 | Predicted triage to surgery in cancer centre RMI≥250 | Total |
| --- | --- | --- | --- | --- | --- | --- | --- |
|  | General Gynae | Cancer Unit | Cancer Centre |  |  |  |  |
| Premenopausal (including perimenopausal) | 16 | 4 | 10 | 30 (9.8%) | 51 | 8 | 59 (19.3%) |
| Postmenopausal | 10 | 2 | 13 | 25 (8.2%) | 43 | 15 | 58 (19.0%) |
| **Total** | 26 | 6 | 23 | 55 (18.0%) | 94 | 23 | 117 (38.2%) |

### Diagnostic test characteristics

RMI at 250-threshold had a sensitivity of 66.7% (95%CI: 39.1%-86.2%%) and specificity of 94.9% (95%CI: 91.8%-96.9%) for OC detection. The positive predictive value (PPV) was 34.8% with negative predictive value (NPV) 96.3%. Should RMI at 200-threshold be used in the postmenopausal cohort, 22.5% (69/306) patients would have undergone surgery (RMI≥200 advised laparotomy under cancer centre team n=16, RMI 0-200 requiring surgical intervention with cancer unit/general gynaecology team n=53).

Using the IOTA-ADNEX model at a 10% threshold, the calculated sensitivity here was 92.3% (95%CI: 66.7% to 98.6%) and specificity 88.7% (95%CI 84.6-91.9%). PPV was 26.7% with NPV of 99.6%.

### Patient process outcomes following IOTA-ADNEX ultrasound

60% (184/306) were discharged within three months and this pathway achieved a 42% (139/334) same day discharge rate. Further CT/MRI imaging was performed in 17% (35/212) of patients with benign IOTA ultrasound findings, 31% (15/49) in the low-intermediate risk group and 89% (40/45) in the high-risk group (Table 3).

**Table 3:** Patient process outcomes by IOTA-ADNEX score.

|  | Discharge within 3 months of referral | Same day discharge | Further surveillance | Further imaging |  |
| --- | --- | --- | --- | --- | --- |
|  |  |  |  | CT | MRI |
| **Benign IOTA findings (N=212)** | 165 (78%) | 126 (59%) | 30 (14%) | 17 (8%) | 18 (9%) |
| **Low-Intermediate risk (N=49)** | 16 (33%) | 5 (10%) | 22 (45%) | 0 (0%) | 15 (31%) |
| **High risk (N=45)** | 3 (7%) | 0 (0%) | 15 (33%) | 22 (49%) | 18 (40%) |

## Qualitative Findings

Five themes were identified from the qualitative analysis.

### Theme 1: Organisational infrastructure

#### Facilitators

Prior experience with OSC models for other gynaecological conditions like postmenopausal bleeding, enabled an established operational framework that could be adapted for this clinic.

Dedicated clinical leadership proved essential as initial absence of designated leads created challenges. Once gynaecologist and sonographer lead roles were established, implementation proceeded more smoothly with improved coordination between services. At SWBH, sonographers valued the support of a lead gynaecology sonographer who oversaw quality assurance and developed an IOTA-reporting proforma to guide scan reporting.

> *‘It helps that … is now Gynae lead for scanning.’*
>
> Sonographer 2

#### Barrier

Sonographers reported that suboptimal imaging quality from current scan machines hindered confident interpretation and could necessitate additional investigations. Therefore, higher-specification ultrasound scanning equipment was required for more accurate reporting. Staffing limitations, including the national sonographer shortage and difficulties in retaining IOTA-certified staff was a recognised restriction on service capacity and skill development as regular IOTA scan exposure was seen as essential for building confidence and expertise. At Walsall, reliance on a single clinician for scanning and assessment limited the number of patients seen and led to cancellations during their absence. Initial underestimation of appointment lengths led to overbooking of patients, causing delays and clinic overrunning.

### Theme 2: Clinical decision making

#### Facilitator

IOTA-ADNEX two-step strategy within a OSC setting enhanced clinical decision-making efficiency. It enables confident triage of benign masses and provided detailed risk stratification to facilitate timely diagnostic decisions and management plan within a single appointment.

> *‘It saves a lot of time in the decision making…It [IOTA-ADNEX in the OSC] facilitates care planning, we can take the decision then and there.’*
>
> Gynae-oncologist 3

This process can help reduce delays in delivering appropriate care for both benign and malignant cases. IOTA-ADNEX assessment was perceived to reduce unnecessary CT/MRI requests, resulting in more efficient resource utilisation.

#### Barrier

Despite the positive clinical impacts, changes take time and can come with resistance. Initial concerns were raised by a clinician of using IOTA-ADNEX rather than RMI as mandated by national guidelines. This was highlighted prior to IOTA-ADNEX being recommended in BGCS guidelines. Early uncertainty around score calculation and interpretation, particularly before the IOTA-ADNEX calculator became readily available, undermined confidence in clinical decision-making.

### Theme 3: Communication and Pathway Definition

#### Barrier

The importance of communication pre-implementation was echoed by clinic staff (nursing staff and health care assistants) who reported insufficient information about this new service.

Clinicians expressed uncertainty in interpreting IOTA-ADNEX scores and requested a clear guidance on discharge, surveillance, further imaging, surgery, and cancer centre referral. Similarly, concerns were raised over the lack of clarity and ownership for those requiring surgery with benign masses and discharged from cancer pathways.

> *‘The patient pathway beyond the OSC needs to be developed’*
>
> Gynaecologist 2

Initial implementation saw most patients referred to multidisciplinary team (MDT) meetings as a “safety net,” creating workload concerns.

#### Facilitator

Establishing post-clinic pathways standardised management based on IOTA-ADNEX scores. Dedicated gynaecology leadership and growing familiarity with the model facilitated appropriate MDT referrals and helped manage the initial rise in MDT workload. Clinician sentiments echoed this concept.

> *‘If it’s working as intended, it should be a better use of resources…more efficient and saving MDT time’*
>
> Gynaecologist 4

### Theme 4: Professional collaboration and training support

#### Barriers

Initial resistance to inter-professional working, particularly regarding sonographers’ expanded diagnostic roles with concerns about quality control and result interpretation were raised.

#### Facilitators

Although resistance to interprofessional working posed a barrier to professional collaboration, interviews identified instances where professionals embraced working together. Real-time case discussion during clinic fostered positive professional relationships and immediate feedback opportunities, helping “professional development and team development.”

Regular multidisciplinary quality assurance sessions proved valuable for continuous learning and development. These sessions, sometimes including an IOTA specialist adviser, enabled case review, image-histology correlation, and continuous skill development.

> *‘To set up a new service like this, it really needs regular meetings between sonographers and gynae-oncologists’*
>
> Sonographer 1

### Theme 5: Patient experience

#### Facilitator

Majority of participants felt that patients benefitted from this model due to reduced waiting times, fewer appointments, and same-visit results discussion. Staff noted that patients appeared “happier and more reassured” and “less anxious” following OSC visits.

#### Barrier

While the service appears beneficial to patients, some raised that this could also be ‘overwhelming’ for some who were not prepared for the possibility of bad news.

This may be related to insufficient pre-appointment patient information. The fast referral to appointment time and absence of clinic-specific information limited the opportunity for adequate patient preparation, particularly for those with lower health literacy and/or language barriers who may have need further support.

## Discussion

This mixed-method study offers a comprehensive evaluation of how IOTA-ADNEX implementation can positively impact care delivery and patient outcomes while exploring key barriers and facilitators in real-world NHS practice. Clinicians involved showed appreciation to the accelerated diagnostic process and data has shown high rates of same-day discharge with no invasive OC cases missed during this study period, supporting NHS Faster Diagnosis Standards.(7) Cancer testing can cause psychological distress, with negative experiences linked to reduced engagement with care.(22) IOTA-ADNEX triage, particularly in an OSC setting, can contribute to reducing patient anxiety and improving experience as highlighted in the qualitative evaluation while improving clinical outcomes. Inadequate standard pelvic ultrasound reporting led to duplicate IOTA-ADNEX scans in secondary care, underscoring the need for consistent triage and reporting protocol. From both a patient and service capacity perspective, a single robust scan is preferable to multiple assessments.

This pilot demonstrated a 20-percentage point reduction in unnecessary, benign procedures compared to RMI threshold using NICE/RCOG protocols. These findings mirror published validation studies when IOTA protocols were performed by expert ultrasound examiners.(23,24) Notably, we demonstrated similar benefits with IOTA-certified, non-expert examiners. This corresponds with the ROCkeTS study, a prospective diagnostic accuracy study involving qualified, level II sonographers, which found IOTA-ADNEX at 10% threshold outperformed RMI in sensitivity.(11) Our results support integrating IOTA-ADNEX into routine clinical practice and reinforce its reproducibility in varied clinical settings (general hospital to specialised cancer centre) and with non-expert ultrasound examiners.

Study funding supported IOTA training and quality assurance sessions, which were critical to successful implementation. Qualitative feedback highlighted the importance of early stakeholder engagement, awareness and education on the IOTA-ADNEX model and clinic set up as well as access to the IOTA-ADNEX calculator.

High rates of conservative management in postmenopausal women, consistent with RCOG guidance for monitoring benign ovarian cysts, show the potential for service optimisation.(8) Long-term studies show that the risk of complications including torsion and malignancy is <1%, reinforcing the safety of conservative management and minimising the need for repeated follow-up.(15) This approach preserves limited NHS imaging capacity whilst improving patient experience by reducing unnecessary intervention. Wider use of the IOTA-ADNEX two-step strategy, supported by structured pathways and standardised reporting proformas, can streamline care, reduce CT/MRI requests and ease MDT workload. At SWBH, regular collaborative sessions involving sonographers and clinicians were valuable for continuous learning and quality assurance, especially sessions with expert input. The evaluation highlights the importance of dedicated leadership in implementing a new diagnostic pathway. A lead IOTA-certified gynaecologist provided clinical oversight, while a lead sonographer ensured supported training and quality assurance. Prior to implementation, clinicians require targeted education, and practical training to feel confident adopting IOTA-ADNEX in routine practice. Practical considerations including patient leaflets, clinic capacity planning, investing in ongoing training and high-quality ultrasound equipment can help support effective implementation.

## Strengths and Limitations

This study’s strengths include its prospective, mixed-method design, evaluating the IOTA-ADNEX implementation in general hospital and specialised cancer centre settings. It combined quantitative clinical outcomes and qualitative insights on the facilitators and barriers to implementation in routine clinical practice using non-expert IOTA certified ultrasound examiners. The qualitative evaluation offers transferable strategies for a wider healthcare context. It involved a range of stakeholders and was led by a clinical research fellow involved in the clinic which offered contextual insight. The evaluation was co-conducted and reviewed by an independent quality improvement consultant to reduce researcher bias. The relatively small sample size, geographic restriction to two West Midland hospitals and use of the OSC model may limit generalisability across the broader NHS where infrastructure and clinical pathways may show greater variation. Retrospective calculation of RMI scores were used to compare outcomes against prospective IOTA-ADNEX triage, which may overestimate differences in intervention rates due to unmeasured factors including patient preferences, comorbidities and surgical fitness. Sensitivity and specificity estimates may be affected by verification bias. During early implementation, the IOTA-SR model was used due to the temporary unavailability of the IOTA-ADNEX calculator and initial limited awareness of the model.

## Conclusion

Implementation of IOTA-ADNEX ultrasound triage within one-stop clinics involving IOTA-certified, non-expert ultrasound examiners represent a valuable quality improvement opportunity in diagnostic pathways. We show that this model can offer improvement in resource capacity, reduction in benign surgery and improved patient experience, especially where expert ultrasound examiners may not be available. However successful implementation of a new diagnostic pathway is an iterative process that requires thoughtful planning around service delivery. This includes establishing clear leadership roles, ensuring access to appropriate ultrasound equipment and providing supportive structures such as defined patient pathways and standardised reporting templates to promote consistency. Ongoing investment in training and effective communication, particularly within the clinical team, is essential to foster strong professional collaboration and sustained adherence.

These findings support wider IOTA implementation across the NHS though further multi-site research is needed to capture variation across hospital trusts. Implementation is underway across 12 hospital trusts within the East of England Cancer Alliance. A larger mixed-method evaluation, the ADVOCATE (ADNEX implementation for Ovarian Cysts Triaging East of England) study, is in progress to assess implementation outcomes across more varied hospital settings and geographies to further validate these promising results.

## Data Availability

All data produced in the present study are available upon reasonable request to the authors

## Patient and Public Involvement

Patients or the public were not involved in the design, or conduct, or reporting, or dissemination plans of our research.

