## Supplementary figures and images for "A mixed-methods evaluation of the implementation of IOTA-ADNEX ultrasound triage in NHS secondary care ovarian diagnostic one-stop clinics"

### supplementary material

***Appendix 1:*** *Current pathway for suspected ovarian cancer*

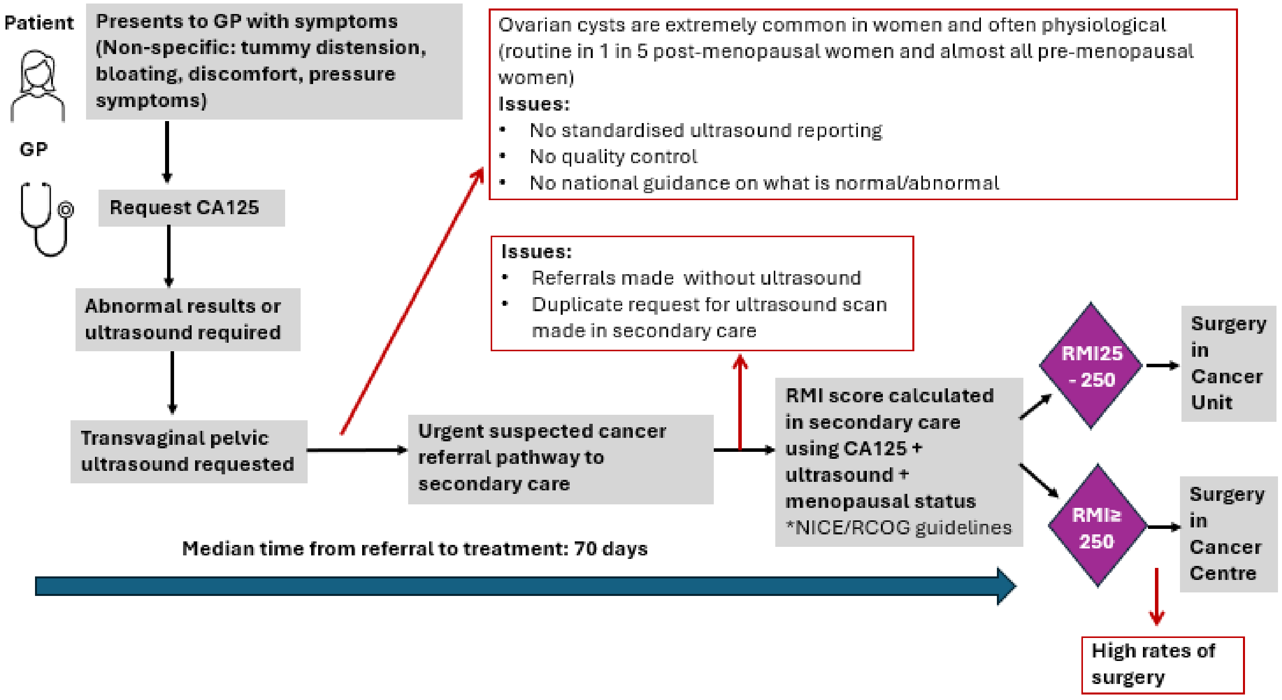
